# Downsides of face masks and possible mitigation strategies: a systematic review and meta-analysis

**DOI:** 10.1101/2020.06.16.20133207

**Authors:** Mina Bakhit, Natalia Krzyzaniak, Anna Mae Scott, Justin Clark, Paul Glasziou, Chris Del Mar

## Abstract

**Objective:** To identify, appraise, and synthesise studies evaluating the downsides of wearing facemasks in any setting. We also discuss potential strategies to mitigate these downsides.

**Methods:** PubMed, Embase, CENTRAL, EuropePMC were searched (inception-18/5/2020), and clinical registries were searched via CENTRAL. We also did forward-backward citation search of the included studies. We included randomised controlled trials and observational studies comparing facemask use to any active intervention or to control. Two author pairs independently screened articles for inclusion, extracted data and assessed the quality of included studies. The primary outcomes were compliance, discomforts, harms, and adverse events of wearing facemasks.

**Findings:** We screened 5471 articles, including 37 (40 references); 11 were meta-analysed. For mask wear adherence, 47% more people wore facemasks in the facemask group compared to control; adherence was significantly higher (26%) in the surgical/medical mask group than in N95/P2 group. The largest number of studies reported on the discomfort and irritation outcome (20-studies); fewest reported on the misuse of masks, and none reported on mask contamination or risk compensation behaviour. Risk of bias was generally high for blinding of participants and personnel and low for attrition and reporting biases.

**Conclusion:** There are insufficient data to quantify all of the adverse effects that might reduce the acceptability, adherence, and effectiveness of face masks. New research on facemasks should assess and report the harms and downsides. Urgent research is also needed on methods and designs to mitigate the downsides of facemask wearing, particularly the assessment of alternatives such as face shields.

## Background

Respiratory viruses can be transmitted by aerosol, droplets and fomites. Facemasks – such as surgical masks, N95 masks, and face shields, and substitutes for surgical masks such as home-made cloth masks – are a physical barrier to aerosol and droplet transmission. During the COVID-19 pandemic, some jurisdictions have implemented policies mandating the use of masks in public places, on public transport or in other crowded environments, to prevent people becoming infected, or infecting others.

While most health organisations mandate the use of facemasks by health workers when caring for patients during a pandemic, recommendations for mask wear in the community vary widely – and include use by all, use only in certain situations (e.g. on public transport, or in crowded places where social distancing is not possible), and no specific recommendations about mask use.

Several trials have evaluated the impact on respiratory infections by use of surgical and N95 masks, which may, at best, modestly reduce acute respiratory infection transmission.^1-3^ Population observational studies suggest that masks have a more substantial effect.^4^ However, the downsides of mask-wearing were either not considered or not reported in most studies.

Anecdotal evidence, and some studies, suggest that there may be a variety of downsides arising from mask use, including: discomfort, sense of difficulty breathing, and communication problems particularly for those who use lip reading. Our aim is to systematically identify and summarise these downsides, to assist policymakers when formulating mask-wearing policies in public settings. We also discuss potential strategies to mitigate downsides of mask-wearing.

## Methods

This systematic review is reported following the Preferred Reporting Items for Systematic Reviews and Meta-Analyses (PRISMA) statement.^5^ We followed the “2 week systematic review” (2weekSR) processes for the review.^6^ The review protocol was developed prospectively (see: https://osf.io/sa6kf/).

### Inclusion criteria

We included studies of people of any age or gender, in any setting. We included studies of any face covering aimed at reducing virus transmission, including surgical masks, N95 masks, cloth masks (both homemade and commercially available). Studies evaluating the use of masks for non-virus transmission purposes were excluded (see Appendix 1 for a complete list of excluded facemasks).

We included studies comparing the use of facemask to any active intervention (e.g. another mask, or another intervention such as hand-washing etc.), and studies comparing the use of facemask to nothing (control) in situations where their use was not mandatory.

We included only primary studies – i.e., randomised controlled trials (RCTs) and observational studies of any design. We excluded studies that could not provide a quantitative estimate of the size or frequency of adverse effects such as case reports, case series, as well as qualitative studies, and reviews.

### Search strategies

We searched PubMed, Embase, Cochrane CENTRAL, EuropePMC (inception-18/5/2020). The search string was designed for PubMed and translated for use in other databases using the Polyglot Search Translator (Appendix 2).^7^ Clinical trial registries were searched via Cochrane CENTRAL, which includes the WHO ICTRP and clinicaltrials.gov. On 22/5/2020, we conducted a backwards and forwards citation analysis in Scopus, on all of the included studies.

No restrictions by language or publication date were imposed. We included publications that were published in full; abstract only publications were included if they had an accompanying record (e.g. trial registry record, or another public report), with additional information.

### Study selection and screening

Four authors (MB&NK, AMS&JC) independently screened the titles and abstracts against the inclusion criteria. One author (JC) retrieved full-text, and four authors (MB&NK, AMS&JC) screened the full-texts. Disagreements were resolved by discussion, or a third author (PG or CDM).

### Data extraction

Data extraction form was piloted on three studies. Four authors (MB&NK, AMS&JC) extracted the data. (See Box 1)

#### Box 1.

List of extracted information

- ***General information:*** study authors, location, study design, duration, aim, setting
- ***Participants:*** health status, disease (if applicable), sample size, age, gender, smoker status, co-morbidities
- ***Intervention and Comparator(s):*** number of participants, type of face covering, adjunct interventions, number of face coverings used, duration of use, disposal
- ***Outcomes:*** definition, measurement instrument, number of adverse events or harms reported (The outcomes were discomforts, harms, and adverse events of wearing face masks Adherence to facemask wearing, Misuse of masks, Discomfort and other physical irritation from masks, Psychological outcomes (e.g. fear, etc.), Dyspnoea (difficulty breathing, shortness of breath) and other physiological impacts, Communication impacts, and mask contamination).

### Assessment of risk of bias in included studies

Four authors (MB&NK, AMS&JC) independently assessed the risk of bias using the Cochrane Risk of Bias Tool.^8^ Disagreements were resolved by discussion or a third author (CDM or PG). Each potential source of bias was graded as low, high or unclear, and judgements were supported by a quote from the study.

### Measurement of effect and data synthesis

Where feasible (≥2 studies reporting the same outcome), we expressed outcome measures as odds ratios (OR) with 95% confidence intervals. Anticipating considerable heterogeneity among the included studies, we used a random effects model. We reported the adherence to facemask wear using risk difference rather than odds ratio, to more clearly convey the differences in adherence between the intervention and control group (not pre-specified in the protocol).

When meta-analysis is not possible or appropriate, we followed the guidance of the Cochrane Collaboration (Cochrane Handbook Section 12.2).^8^ When narrative synthesis was required, we reported the results separately for each harm or adverse outcome.

The individual was used as the unit of analysis, where possible; otherwise, we extracted the information as it was presented, e.g. the number of harms in each group. We attempted to contact investigators or study sponsors to provide missing data.

We used the I^2^ statistic to measure heterogeneity. Because we included fewer than 10 trials, we did not create a funnel plot.

We did not pre-specify subgroup or sensitivity analyses. However, as data was available, we conducted a subgroup analysis of adherence to mask-wearing by studies which evaluated facemask wear alone, and those evaluating facemask together with handwashing.

## Results

Database searches identified 4691 publications, supplemented with 2035 references from forward and backward citation searches and other sources, totalling 6726. After de-duplication, 5471 references were screened by title and abstract; we full-text screened 214 references, excluding 174 (see Appendix 3 for a list of excluded studies with reasons). We included 40 articles ^9-48^ corresponding to 37 studies (Table 1 reports the characteristics of included studies), and meta-analysed 11 studies.^12,18,19,23,28-31,38,42,44^ (Figure 1).

**Table 1.**
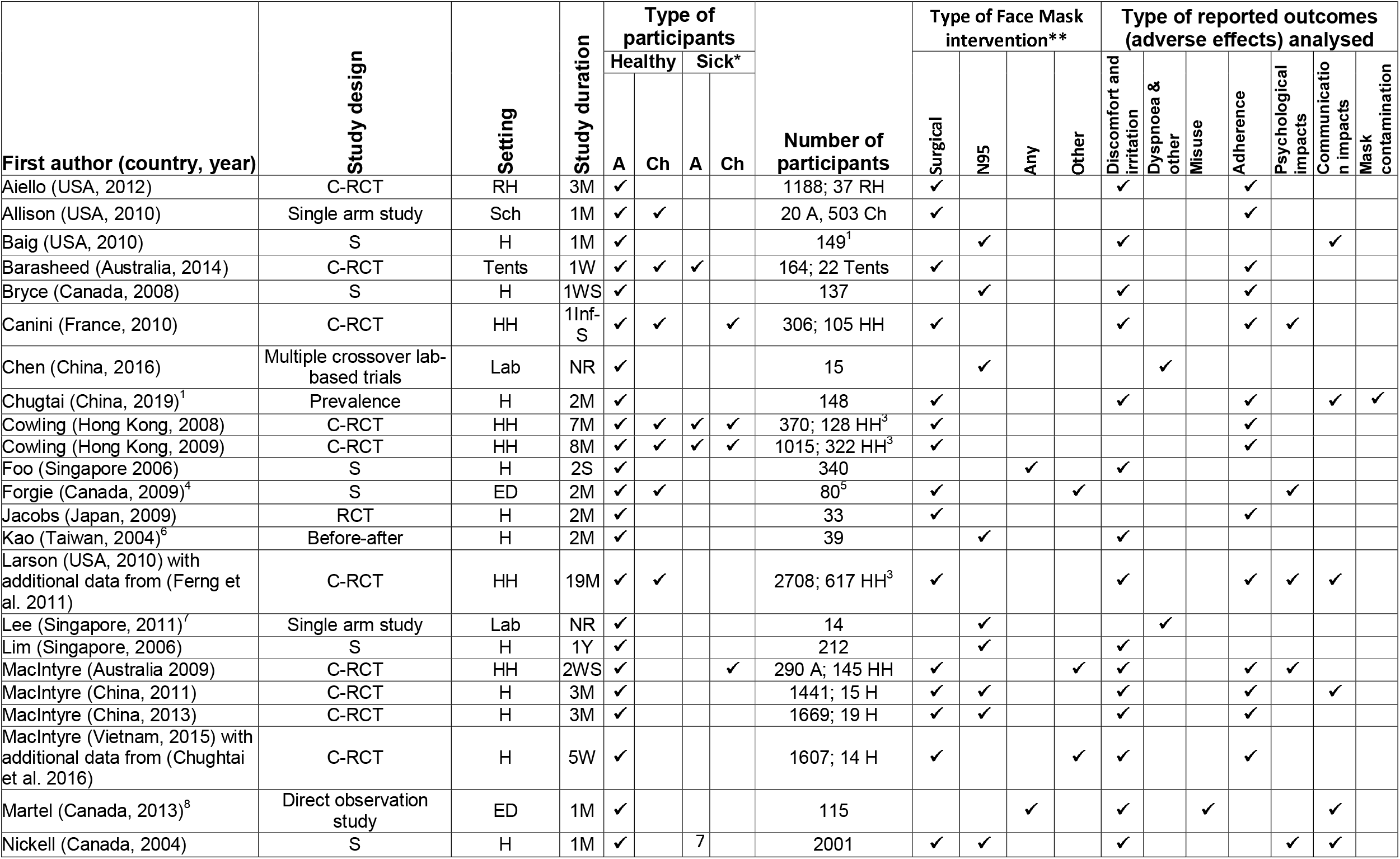
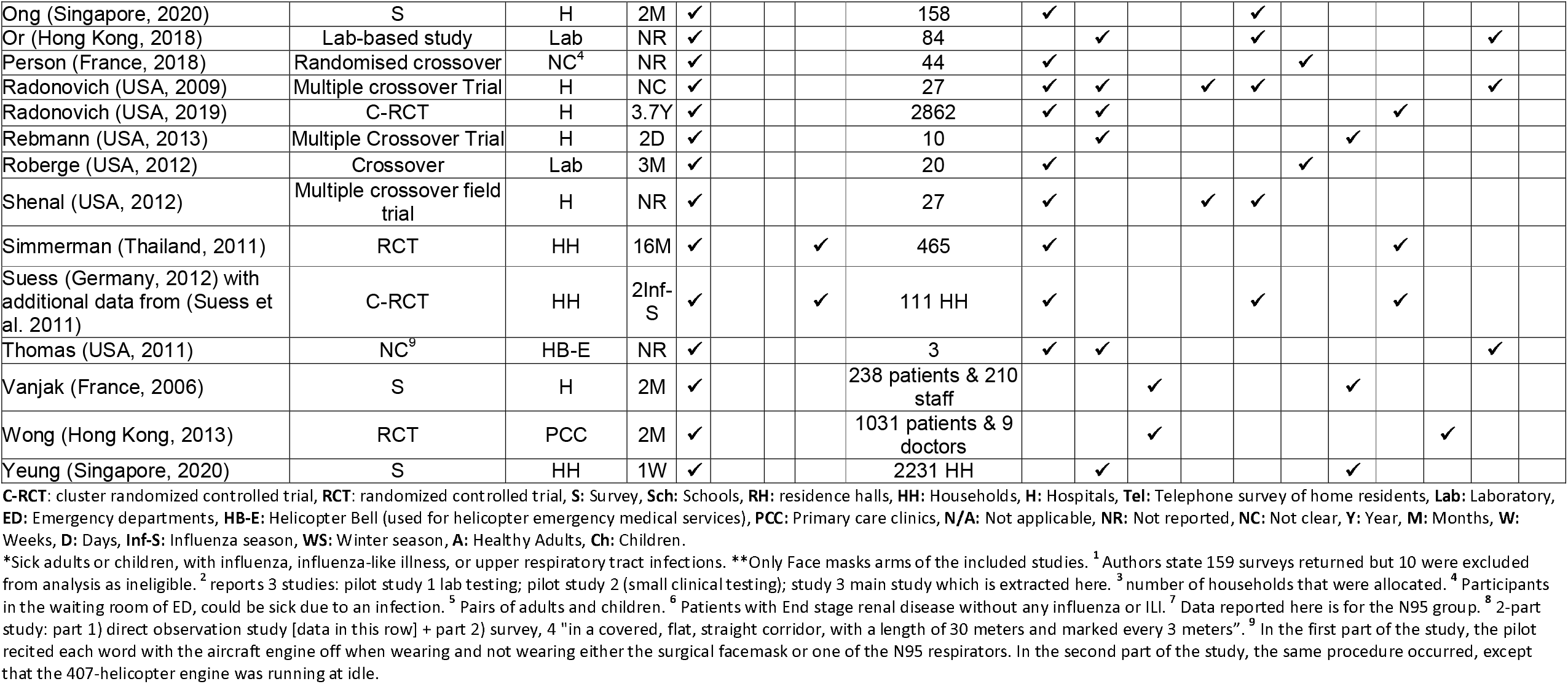
Characteristics of included studies.

**Figure 1.**
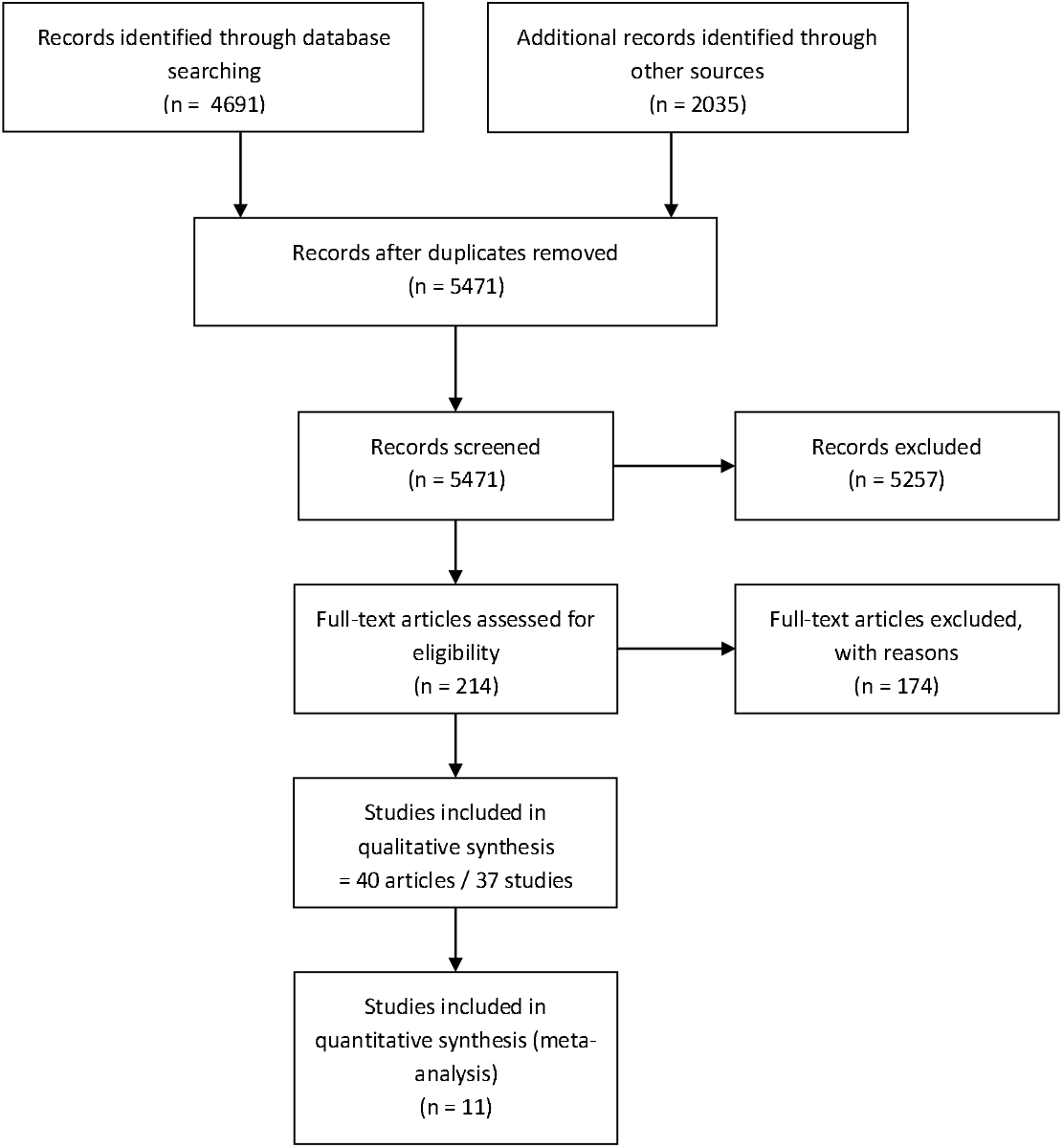
PRISMA flow diagram

### Risk of bias

Inclusion of both observational studies and RCTs could explain the high risk of bias across the included studies. Reporting of sequence generation, allocation concealment and blinding of outcome assessment was poor in 20%-30% of studies. Due to the nature of interventions, blinding of participants was rare. For RCTs, we reported blinding of outcome assessment for the main trial (not the adverse events) – as some outcomes were lab-confirmed and were considered of low risk. We found no evidence of incomplete outcome data or selective reporting of outcomes. Funding statement, funder’s role and authors’ conflict of interests were adequately reported in most studies (see figure 2, and figure 3 in Appendix 4).

**Figure 2.**
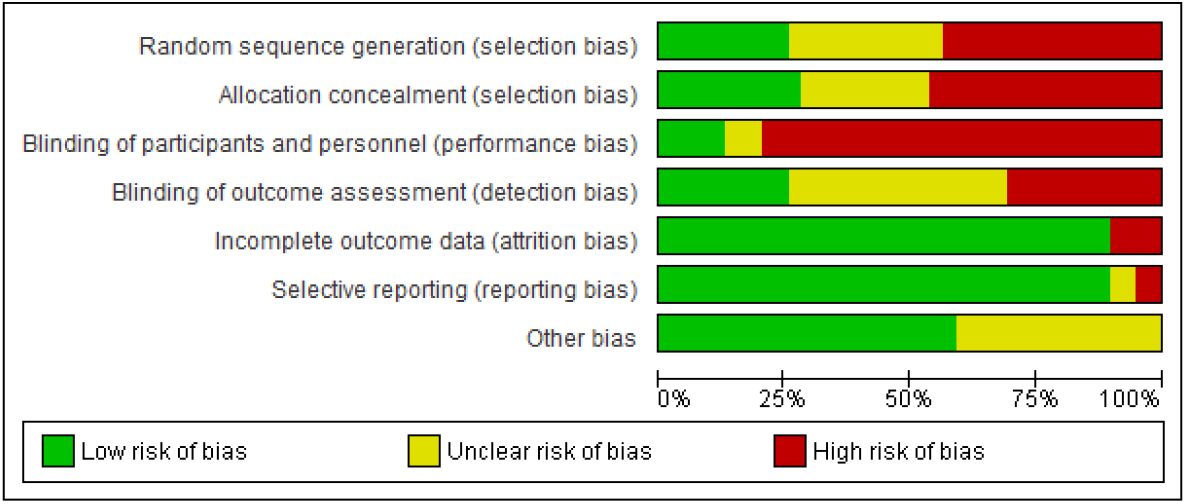
Risk of bias graph: review authors’ judgements about each risk of bias item presented as percentages across all included studies.

**Figure 3.**
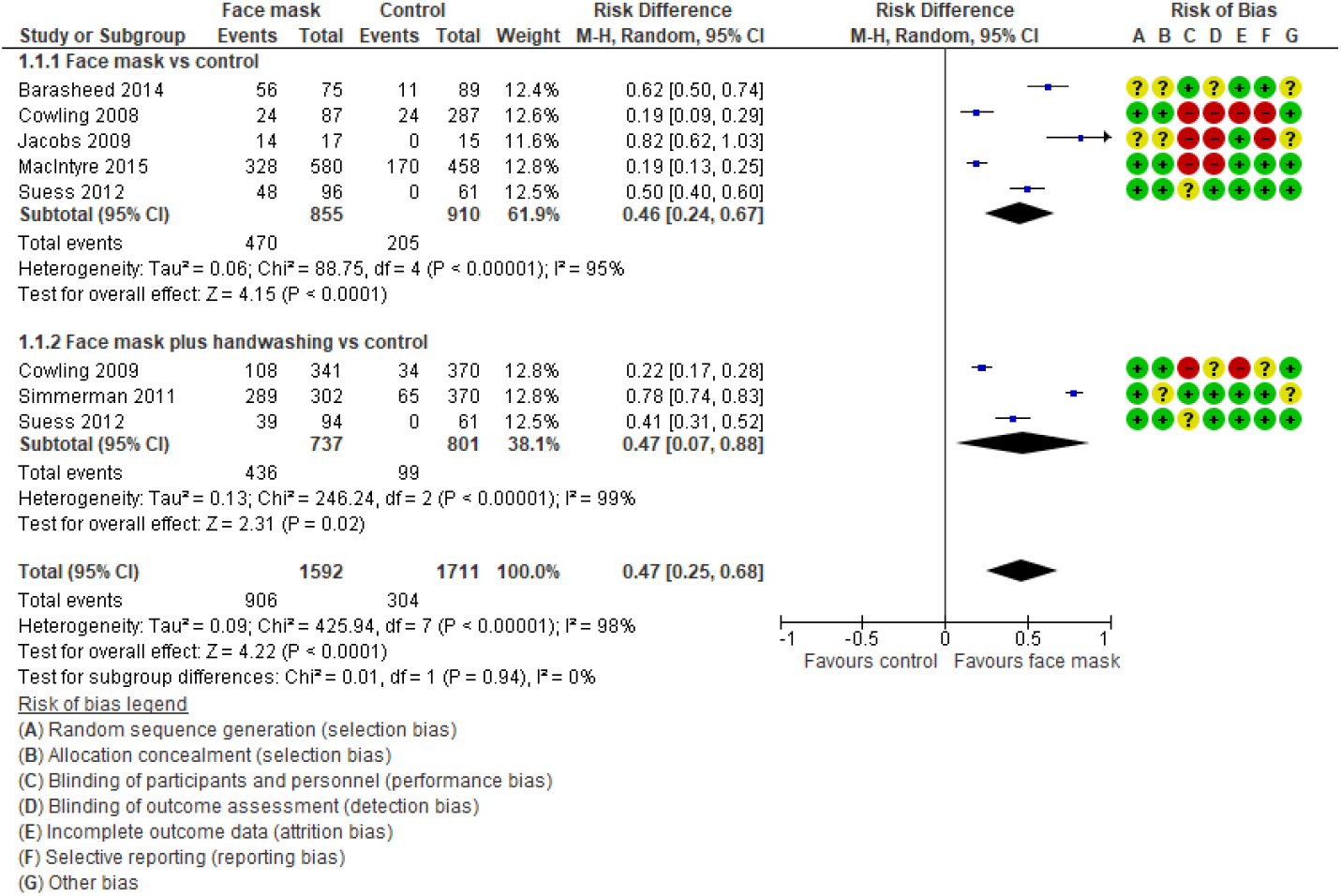
Comparison of adherence to face masks vs control

### Adherence to facemask wearing

17 studies (14 RCTs^9,12,14,18,19,23,25,28-31,38,42,44^, 3 observational^10,13,17^) reported on adherence to facemask wearing; 11 were meta-analysed.

#### Face mask versus control comparison

Comparison of face masks to control was subgrouped into studies comparing facemask alone vs control; and studies of facemask plus handwashing vs control. (Figure 3)

Facemasks alone vs control (5 studies) showed the the facemask group had a significantly higher facemask wear compared to control (Risk Difference (RD): 0. 46, 95% CI 0. 24 to 0. 67, P < 0.0001). Studies evaluating facemask plus handwashing vs control (n=3), similarly showed significantly higher facemask wear in the facemask group (RD: 0. 47 (95% CI 0. 07 to 0. 88, P < 0. 0001). Overall, 7 studies (3303 participants) compared facemasks to control. Facemask wear was 47% higher in the facemask group, although heterogeneity was very high (RD: 0. 47, 95% CI 0. 25 to 0. 68, P < 0. 0001, I^2^ = 98%.)

We explored the possible sources of heterogeneity. Excluding studies with 3 or more domains at high risk of bias did not decrease heterogeneity (I^2^ = 96% for facemask vs control; 99% for facemask plus handwashing versus control) (Figure 4, Appendix 4). We excluded study population as the source of heterogeneity, because subgrouping studies by those in a community/household settings (which included both index cases and their contacts) versus those in a hospital setting (which included healthy healthcare workers) likewise did not decrease heterogeneity (I^2^ = 99% for community/household studies, and 97% for hospital studies) (Figure 5, Appendix 4). We excluded intervention and control as sources of heterogeneity, since all studies compared medical/surgical masks to control (no mask) – although some mask-wear did occur in the control groups.

**Figure 4.**
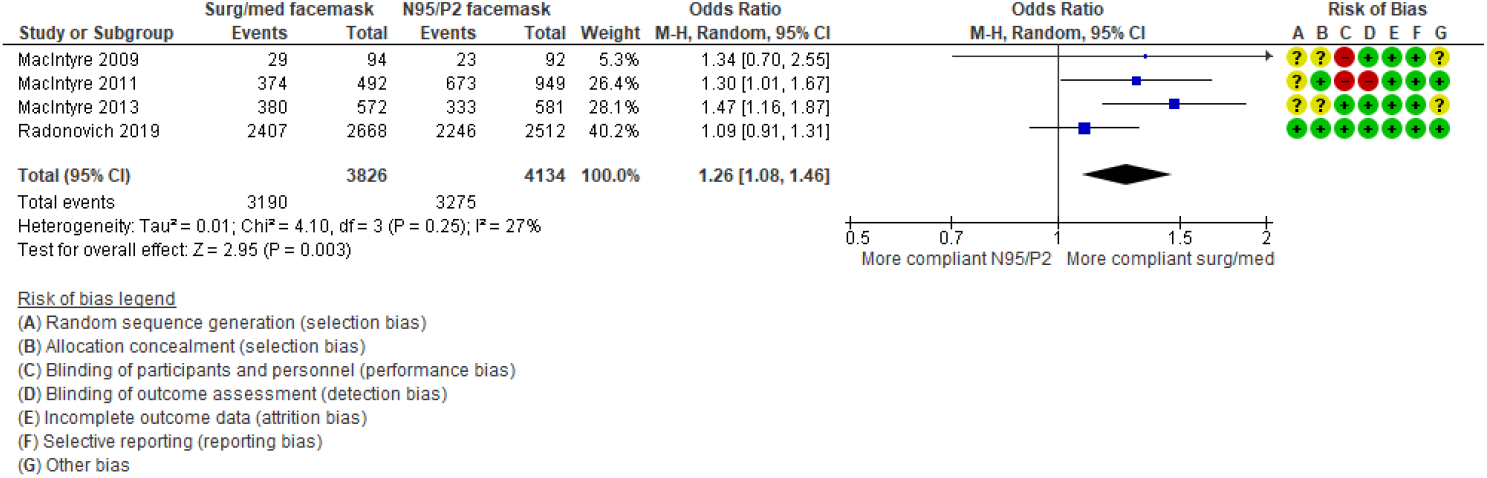
Comparison of adherence to surgical/medical face masks vs N95/P2 masks

We consider the outcome measurement to be the most likely other source of heterogeneity. All studies relied on self-reporting of the outcome; only one verified this by counting the number of masks used.^18^ What was considered ‘wearing a facemask’ varied: it was unclear,^12,18,42^ involved wearing a facemask “always or mostly/often”,^19,44^ included wearing a facemask whilst on hospital property, performing a healthcare worker role,^23^ and included facemasks wear during shift for 70% of time or more.^29^ The follow-up was very short (5-9 days) for 4 studies;^12,18,19,44^ it was longer for 3 studies (21-77 days).^23,29,42^

#### Facemask (surgical/medical) vs facemask (N92/P2 mask)

Four studies (7960 participants) compared adherence for different facemasks.^28,30,31,38^ Facemask wear was significantly higher in the surgical/medical facemask group than in the N95/P2 group, OR 1.26 (95% CI 1. 08 to 1. 46, P < 0. 01). Heterogeneity was very low (I^2^ = 27%). (Figure 4)

### Studies not included in the meta-analysis

#### RCTs (n=3)

One study reported similar duration of facemask wear per day in the facemask alone group vs facemask plus handwashing group (mean of 5.08 and 5.04 hours/day, respectively).^9^ Another reported that within the facemask group, there were no significant differences between individuals with ILI among contacts vs no ILI among contacts, for facemask use.^14^ Finally, 22 of 44 households randomised to the ‘education with sanitiser and facemasks’ arm, reported having used a mask within 48 hours of episode onset.^25^

#### Observational studies (n=3)

In elementary school setting, approximately twice as many teachers as students wore face masks.^10^ A mean compliance score with N95 use guidelines was 21.2 (on a 25-point scale) among frequent users of N95 respirators in a hospital setting.^13^ Another study found that majority of survey respondents (91%) wore 1-2 masks per day (range 1-4).^17^

### Misuse

Mask misuse appears less studied than other harms and discomforts. A study of 10 nurses observed for 10 minutes/hour over 2 shifts found that they touched their face 2-3 times per hour, their mask 5 times per hour, and their eyes once per 2 hours, when observed by students.^39^ In a study of health workers, 13 of the 53 who responded (25%), reported wearing masks only covering their mouth, not their nose.^46^ A study in two hospitals,^32^ observed triage nurse behaviour with 118 patients with fever and cough, found that in only 18% of cases the nurses informed patients of the need to wear mask, and in half of those, gave instruction on the need to cover both mouth and nose. A cross-sectional study evaluating the proficiency of the Singaporean public in wearing N95 masks found only 90/714 subjects passed the visual mask fit test; the most common criteria performed incorrectly were: strap placement, leaving a visible gap between the mask and skin, and tightening the nose-clip.^48^

### Discomfort and irritation

Several RCTs of specifically measured mask wear discomfort,^14,28-30,44^ but most only recorded spontaneously reported events ^9,25,31^ or did not report any.^12,18,19,23,38,42,47^ A trial of index influenza cases allocated to wear masks or no mask, found the 51 allocated to masks wore them on average 3.8 hours/day and 38 (76%) reported discomfort (Table 2).^14^ A study of health care workers in Beijing asked to wear masks for their full shift, found 84% complained of at least one problem (Table 2).^17^ In a German study, 65/172 participants reported problems with mask wearing – most commonly warmth, pain and shortness of breath.^44^

**Table 2.** Types of discomfort assessed in trials of face masks used to prevent viral transmission.

| First author, year<br>(Type of mask) | Population,<br>Number | Difficulty<br>Breathing | Facial<br>irritation or<br>discomfort | Headache | Other |
| --- | --- | --- | --- | --- | --- |
| Canini, 2010<sup>14</sup><br>(surgical masks) | Household, 105<br>index cases | 34% | 14% | - | 46% warmth |
| Chughtai 2019<sup>17</sup><br>(surgical masks) | Health care<br>workers, 148 | 12% | 17% | 6% | - |
| MacIntyre 2011<sup>30</sup><br>(Surgical) | Health Care<br>workers, 492 | 12% | 11% | 4% | - |
| MacIntyre 2011<sup>30</sup><br>(N95) | Health Care<br>workers, 949 | 19% | 52% | 13% | - |
| MacIntyre, 2015<sup>29</sup><br>(Cloth v Medical) | Health Care<br>workers, 1130 | 18% | 35% | - | - |

In a trial of healthcare workers comparing surgical and N95 masks to prevent influenza, more workers found the N95 uncomfortable (42%) than the medical mask (10%) when worn an average of 5 hours per day,^30^ with significant differences in headaches, difficulty breathing, and pressure on the nose (Table 2). A trial of cloth versus medical masks in healthcare workers found similar rates of discomfort.^29^ A community trial comparing surgical and P2 (N95) masks found >50% reporting concerns, primarily discomfort, with similar rates (15% vs 17%) across groups.^28^

Discomfort increases with duration of mask wearing. A crossover field trial of 27 healthcare workers found increased discomfort over time; half the subjects were unwilling to wear a medical mask for the full 8-hour shift despite regularly wearing them for short periods.^41^

Two surveys of healthcare staff in Singapore during the SARS epidemic assessed headache and skin reactions.^21,27^ In one, 79/212 (37%) reported face-mask-associated headaches, 26 (33%) reported headache frequency exceeding six times/month, and 6 had taken sick leave.^27^ Another survey of healthcare workers in Singapore, found that of the 307 staff who used masks regularly, 60% reported acne, 51% facial itch, and 36% rash from N95 mask use.^21^ A COVID-19 survey of healthcare workers in Singapore found that 128/158 (81%) developed *de novo* PPE-associated headaches, increasing with duration of use (> 4 hours).^34^

One study (2×2 factorial design) examined the potential of mask fit test and training to mitigate discomfort, but found no clinically or statistically important differences between arms.^35^

Six observational studies reported either general discomfort^33^ or spontaneously reported events among participants who wore facemasks.^11,13,24,32,37^

### Psychological

6 studies reported on psychological impacts from wearing face masks (4 RCTs, 2 observational).

#### Fear

A 3-arm RCT found significantly higher risk perception scores in the mask group (38/60) than non-mask groups (30/60); participants in the mask group were more fearful that they and their family would get sick from the flu.^20,25^

In an observational study, children in a paediatric emergency department waiting room (n= 80) were shown pictures of clinicians wearing either a surgical mask or a clear face shield; 18 children (22.5%) reported surgical masks to be more frightening due to an inability to see clinicians faces, and 14 children (17.5%) reported face shields to be more frightening. However, 47 children (59%) reported that neither were frightening.^22^

#### Stigma

In a 2-arm cluster-RCT, 15 (29%) patients wearing masks reported they did not like being seen wearing a mask.^14^ In a 3-arm RCT, more children reported disliking their parents wearing a P2 mask than a surgical mask (8/92 vs 6/94), however the difference was not significant.^28^

#### Loneliness

One observational study reported on the loneliness outcome. In a survey investigating the psychosocial effects associated with working in a hospital during the SARS outbreak, 222 (13%) respondents reported a sense of isolation as one reason masks were perceived as bothersome.^33^

#### Empathy

One RCT reported that the wearing of a face mask by doctors had a negative effect on patient’ perceptions of the doctors’ empathy during consultations, with a mean Consultation and Relational Empathy (CARE) score in the mask group of 33.93 (SD= 7.65), and 34.91 (SD=7.84) in the no mask group.^47^

### Dyspnoea and other physiological consequences

Studies of physiological impacts were generally done on masks designed for dust, vapours, and other non-transmission purposes; few studied surgical or N95 masks.

A French crossover study (44 subjects) found surgical masks had no impact on 6-minute walking time, but subjects had an increased sense of dyspneal with a mask: 5.6 vs 4.6cm on a 10cm visual analogue scale (p<0.001),^36^ which may come from the increased effort required. A study in 14 adults found that N95 masks increased respiratory resistance in 30 seconds of breathing by over 100%, resulting in average reduction in nasal spirometry of 37%.^26^ A study of 20 subjects on a treadmill found the surgical masks increased respiratory rate by 1.6 breaths/minute (p=0.02), heart rate by 9.5 beats/minute (p<0.001), and transcutaneous CO2 levels of 2.2 mmHg (p<0.001).^40^ Finally, a Chinese study of 15 subjects wearing a monitoring garment for respiratory signals found that N95 masks increased both subjective breathing resistance (from none to mild) and increased respiratory rate, the muscle activity of sternomastoid, scalene, diaphragm and abdominal, the fatigue of scalene and intercostal.^15^

### Communication

Nine studies (2 RCT, 7 observational) reported on communication difficulties while wearing facemasks.

A trial comparing the use of surgical and N95 masks by healthcare workers to prevent influenza, found more workers in the N95 mask group than the surgical mask group reported mask causing trouble with patient communication (8% vs 3%).^30^ Another trial of 15 participants who wore a surgical mask for approximately 60 minutes while performing various tasks,^20,25^ found that participants did not report any interference with communication while answering the phone. In a cross-over trial of 27 healthcare workers,^37^ more participants in the surgical mask group reported diminished communication acuity (visual, auditory or vocal) as the reason for discontinuing mask use before the end of an 8 hour shift (7 complaints compared to 4 complaints among N95 mask wearers).

Of 2001 healthcare workers in Toronto responding to a survey during the SARS outbreak, difficulty communicating (47%), and difficulty recognizing people (24%) were identified as key reasons masks (surgical or N95) were perceived as being particularly bothersome.^33^ In a survey of 149 healthcare workers,^11^ 41 (27.5%) of respondents reported a difficulty ‘always’/’most of time’ in verbally communicating with patients while wearing a mask.

In another Canadian survey (115 healthcare workers),^32^ 26 (23%) respondents reported that wearing masks interfered with their relationships with their patients. Among 148 healthcare workers asked to wear a mask during a 6-8 hour shift, 11 (7.4%) reported trouble communicating with patients.^17^

In a study of 3 participants evaluating the impact of wearing a surgical or N95 mask on radio reception, all participants were able to accurately record all pilot-recited words regardless of the type of mask worn by the pilot. However, when the aircraft engine was turned on, the accuracy decreased for the N95 mask, compared to surgical or no mask.^45^

In another study, the performance or absence of fit testing prior to mask use did not affect communication, as 2 participants (out of 21) in each group reported ease of talking to be unsatisfactory.^35^

### Mask contamination and other issues

One concern about mask use is the potential for contamination of the mask surface, and subsequent self-inoculation to the wearer’s eyes or when demasking. No studies examined that directly, but one study of the health care workers found on average 10% of masks had viral contamination after usage, and that was higher for masks worn > 6 hours (OR 7.9) or > 25 patients seen (OR 5.0).^17^ Given the rates of misuse (see Misuse section above) this contamination raises concerns about self-inoculation.

Several authors have raised concerns about “risk compensation” – non-adherence to other precautions because of the sense of protection – but we found no studies that quantify its extent.

## Discussion

We identified 37 studies reporting downsides, harms and adverse events associated with the wearing of facemasks – 15 RCTs and 22 observational studies. The largest number of studies reported on the discomfort and irritation outcome (20 studies), fewest on misuse of mask (4 studies), with no studies directly investigating or quantifying mask contamination or risk compensation behaviour. The only meta-analysable outcome was adherence to facemask wear (17 studies, 11 meta-analysed). 47% more people wore facemasks in the facemask group compared to control, although the percentage of people wearing facemasks in the control group was non-zero in 5 studies; facemask wear adherence was also significantly higher (26%) in the surgical/medical mask group than the N95/P2 group. Risk of bias was generally high for blinding of participants and personnel, and selection bias, and low for attrition and reporting biases.

The review’s strength lies in its inclusion of non-randomised study designs in addition to RCTs, as trials frequently underreport or fail to report harms.^49^ Additionally, the inclusion and exclusion criteria were tested and refined on a test library of 98 references, prior to screening the full search results. The key limitation includes the hospital-setting of most of the included studies: as hospital workers are accustomed to wearing masks, the conclusions may not be fully generalisable to the community. Although this varies among the studies that reported mask use in hospital setting, as there are different confounding factors that may contribute to increased reporting of irrigation (e.g. length of shift, air-conditioning on the wards and whether the staff were wearing the full PPE which adds to the full discomfort). We report two differences between the protocol and the review: first, the comparison of facemask to control in the adherence outcome was reported using risk difference (rather than pre-planned odds ratio) to more clearly convey the differences between the two groups (odds ratio for compliance with facemask wear was reported for the facemask versus facemask comparison, however). Second, not having anticipated data availability, we did not prespecify a subgroup analysis of the intervention (facemask wearing) by studies which evaluated facemask wear alone, and studies evaluating facemask with handwashing.

Several recent systematic reviews have focused on the effectiveness of masks in preventing or reducing viral transmission; some of these reviews reported on harms in the included studies.^5,15,39,50^ However, none specifically focused on the wider set of studies examining the physiological, psychological and other adverse effects addressed in this review. The Cochrane review on physical barriers noted the impact of masks on discomfort and communication in some of the randomised trials, and its findings are consistent with this review, but did not extend to studies with outcomes other than viral transmission or non-randomised study design.^3^

The downsides identified in this review should aid in designing strategies to mitigate problems, and guide the situations where the benefits of masks might outweigh the downsides. As suggested by the higher adherence to surgical masks than to the N95 masks, mitigation of discomforts may also increase adherence to facemask wear, and hence their effectiveness. Mitigation might be achieved by considering of the when, where, and how of mask wearing or by mask redesign or substitution with alternatives (e.g. face shields).

### Limiting circumstances

Use of facemasks should be restricted to higher risk circumstances, including crowded, indoor spaces, where physical distancing is not possible, e.g. public transport. Conversely, exercising outdoors is both low risk and has higher downside of wearing masks, because of the increased perceived dyspnoea.

### Limiting duration of facemask wear

Duration increases both discomfort and non-adherence. Duration might be decreased by demasking during breaks or scheduling mask breaks. Changing masks more often will help with adherence and the contamination risks, but will increase costs and environmental problems with waste disposal.

### Modification for specific groups

Some groups are likely to have greater difficulty with mask wearing adherence and correct usage – including children, some patients with mental illnesses, those with cognitive impairment, or respiratory disorders such as asthma or chronic airways disease, and patients with recent facial trauma or oromaxillofacial surgery.

### Substitution

Face shields provide an alternative to facemasks, which mitigates several of the downsides (e.g. reducing the communication difficulties and breathing resistance), while also provides eye protection. However, there is little evidence on the discomforts of wearing face shields, and on the degree of protection provided. Other innovative mask designs currently being developed, require discomfort and adherence evaluations in addition to the droplet penetration.

Currently, there are insufficient data to quantify all of the adverse effects that might reduce the acceptability, adherence, and effectiveness of facemasks. Any new research on facemasks should also assess and report the harms and downsides. Urgent research is also needed on methods and designs to mitigate downsides of facemask wearing, particularly assessment of alternatives such as face shields.

## Data Availability

No further data available

## Acknowledgements

Our thanks to Tom Jefferson for stimulating this review and providing comments, and to John Conly and Marylouise McClaws for comments on the draft.

## Declarations

### Funding

The present systematic review was conducted as part of the work of the Centre of Research Excellence in Minimising Antibiotic Resistance in the Community (CRE-MARC), funded by the National Health and Medical Research Council (NHMRC), Australia (grant reference number: GNT1153299). The funder had no involvement in this systematic review.

## Notes

### Competing Interest Statement

The authors have declared no competing interest.

### Author Declarations

No ethics approval required

